# Plasma TDP-43 is a potential biomarker for advanced limbic-predominant age-related TDP-43 encephalopathy neuropathologic change

**DOI:** 10.1101/2025.07.09.25331146

**Authors:** Jijing Wang, Julie A. Schneider, David A. Bennett, Nicholas T. Seyfried, Tracy L. Young-Pearse, Hyun-Sik Yang

**Author notes:** Correspondence to: Hyun-Sik Yang, MD.

## Abstract

Limbic-predominant age-related TDP-43 encephalopathy neuropathologic change (LATE-NC) is a common cause of late-onset dementia that does not yet have specific in vivo biomarkers. Here, we examine the biomarker potential of plasma TDP-43, measured using a highly sensitive Nucleic Acid Linked Immuno-Sandwich Assay (NULISA), in detecting advanced LATE-NC. Leveraging plasma TDP-43 and phospho-TDP-43 data from 50 deceased Religious Orders Study and the Rush Memory and Aging Project participants, we show that plasma TDP-43 and pTDP-43 were associated with advanced LATE-NC, especially in those with comorbid Alzheimer’s disease neuropathologic change (ADNC): in the subgroup with autopsy-confirmed AD dementia (n=32), receiver operating characteristic area under the curve (AUC) for both plasma biomarkers approached 0.8. Plasma TDP-43 was also elevated in hippocampal sclerosis, a pathologic finding closely related to advanced LATE-NC. Together, our findings suggest the potential utility of plasma TDP-43 and pTDP-43 as biomarkers of LATE-NC, particularly in individuals with comorbid AD.

## Main Text

Limbic-predominant age-related TDP-43 encephalopathy neuropathologic change (LATE-NC), characterized by TDP-43 (Transactive response DNA binding protein of 43 kDa) proteinopathy predominantly affecting the medial temporal lobe, is a major cause of late-onset amnestic dementia [1]. Currently, no *in vivo* diagnostic biomarker is available. The recently published clinical criteria operationally defined “probable LATE” in older individuals with negative AD biomarkers [2]. However, most LATE-NC cases are comorbid with Alzheimer’s disease neuropathologic change (ADNC), and in vivo diagnosis of LATE-NC remains challenging without a specific molecular biomarker [3]. Prior plasma biomarker investigations of TDP-43 proteinopathies have been limited by detection sensitivity [4]. Measuring peripheral extracellular vesicles (EV) TDP-43 has shown initial promise [5], but EV-based methods are currently limited.

Here, we examine the biomarker potential of plasma TDP-43 measured with a highly sensitive Nucleic Acid Linked Immuno-Sandwich Assay (NULISA) [6] in detecting advanced LATE-NC. We analyzed the data from fifty deceased participants from two extensively characterized clinical-pathologic cohorts—the Religious Orders Study and the Rush Memory and Aging Project [7] (ROSMAP; N = 18 no AD pathology/cognitively unimpaired controls, N = 32 pathology-confirmed AD dementia; **Supplementary Table**)—who also had plasma NULISA assay through a previous study [8]. In brief, LATE-NC stages were evaluated with immunohistochemistry (targeting phosphorylated TDP-43 at Ser409/Ser410; TAR5P-1D3) and documented as follows: stage 0 = no TDP-43 proteinopathy, stage 1 = amygdala only, stage 2 = extension to hippocampus/entorhinal cortex, and stage 3 = extension to neocortex. The quantitative burden of amyloid-beta (Aβ) and paired helical filament (PHF) tau in the brain was measured by immunohistochemistry in eight regions (hippocampus, entorhinal cortex, midfrontal cortex, inferior temporal cortex, angular gyrus, calcarine cortex, anterior cingulate cortex, and superior frontal cortex) and their quantitative burden measures were averaged. Lewy body pathology (present/absent) was assessed with α-synuclein immunohistochemistry. Hippocampal sclerosis (HS) was recorded as present if there was severe neuronal loss and gliosis in the CA1 sector and/or subiculum. Plasma samples proximate to death (average 3.8±1.9 years before death) were analyzed using the NULISA CNS Disease Panel (Alamar Biosciences), following the manufacturer’s protocols. Data were normalized to internal control and inter-plate control values, then log2-transformed to produce NULISA Protein Quantification (NPQ) units. Total TDP-43 and TDP-43 phosphorylated at Ser409 (pTDP43) were used for subsequent analyses. We used Wilcoxon rank-sum tests, Spearman correlation, and receiver operating characteristic (ROC) analyses to assess plasma biomarker–pathology association. We ruled out confounder-driven results by adjusting for demographic covariates (age at death, sex, and time lag between plasma collection and death; adjusted one covariate at a time, given the limited sample size) or tau burden. All study protocols and procedures were approved by the Institutional Review Board at Rush University Medical Center, and all participants signed written informed consent and the Anatomical Gift Act [7].

Plasma TDP-43 and pTDP-43 were associated with advanced LATE-NC, especially in those with comorbid ADNC. We first analyzed the data from all participants (N = 50). Plasma TDP-43 was elevated in individuals with advanced LATE-NC (stage 2 or 3; Wilcoxon W = 202, P = 0.035; **Figure 1A**); pTDP-43 was not (P > 0.05; **Figure 1B**). Both plasma markers showed modest discriminative power in detecting advanced stages of LATE-NC (TDP-43: AUC = 0.68; pTDP-43: AUC = 0.61). Then, we focused on the AD dementia subgroup (N = 32). Both plasma TDP-43 and pTDP-43 were elevated in advanced LATE-NC in this subgroup (P = 6.0×10^−3^ and P = 0.019, respectively; **Figure 1C-D**) and remained so after adjusting for demographic covariates. The ROC AUCs for both analytes approached 0.8 (**Figure 1E**). In the AD dementia subgroup, either plasma marker was associated with Aβ or Lewy body pathology (P > 0.05), supporting the specificity of plasma TDP-43 in detecting LATE-NC. Intriguingly, both markers were inversely associated with tau burden (AD dementia subgroup; TDP-43: Spearman’s rho = -0.51, P = 3.3×10^−3^; pTDP-43: rho = -0.47, P = 7.3×10^−3^); nonetheless, the association of plasma TDP-43 or pTDP-43 with LATE-NC remained significant after adjusting for tau. Notably, plasma TDP-43 was elevated in individuals with hippocampal sclerosis (HS; all participants except one with non-missing data [N =49], W = 36, P = 0.048; **Figure 1F**), whereas plasma pTDP-43 was not.

**Figure 1.**
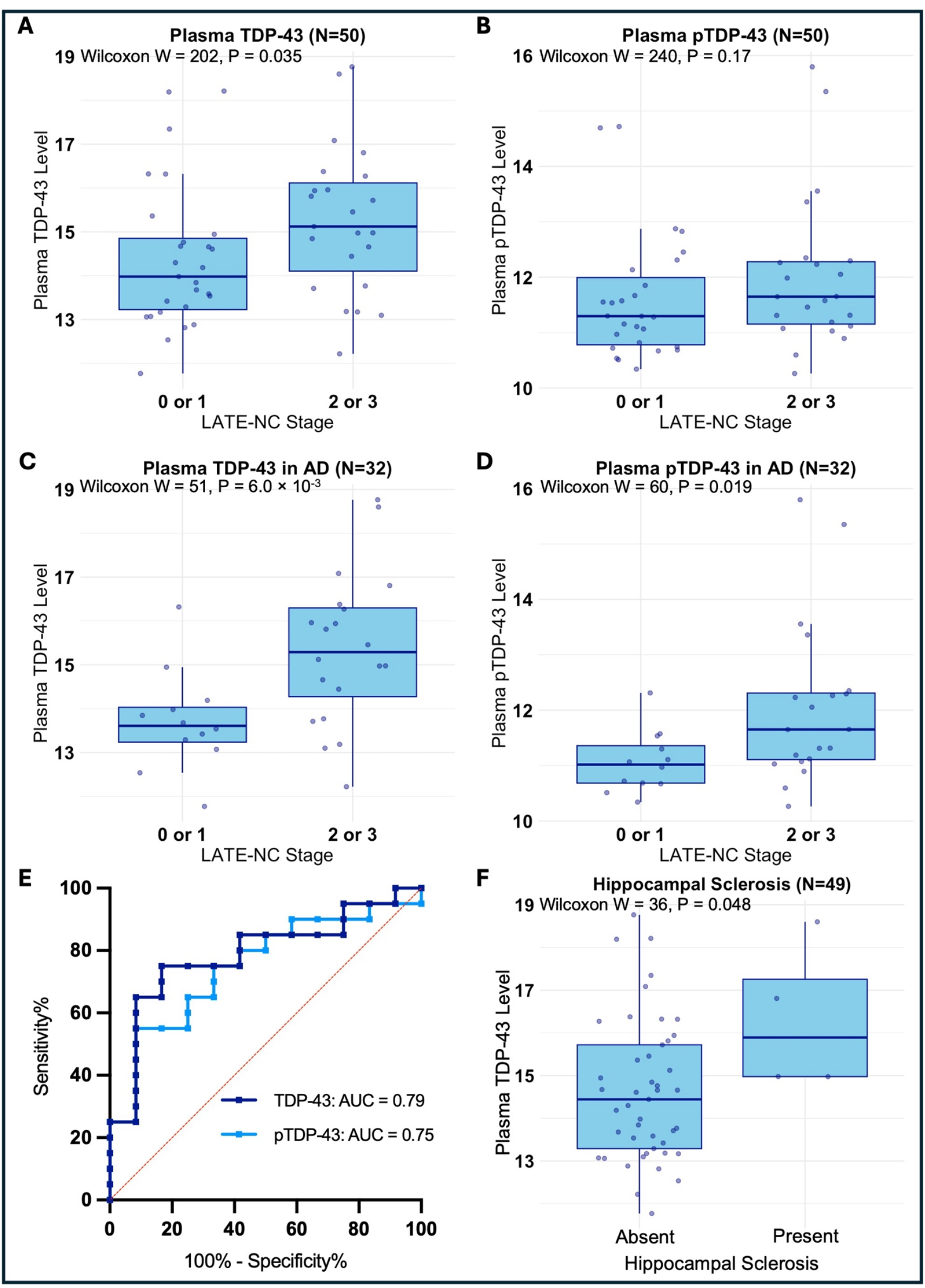
Plasma TARDBP, pTDP-43 (pS409), and post-mortem LATE-NC. **A**, Plasma TDP-43 levels were significantly elevated in advanced LATE-NC. **B**, Plasma pTDP-43 levels were not significantly elevated in advanced LATE-NC. **C-D**, In a subset of individuals with comorbid AD, (**C**) plasma TDP-43, and (**D**) pTDP-43 levels were significantly elevated in advanced LATE-NC. **E**, ROC curves of plasma TDP-43 and pTDP-43 in detecting advanced LATE-NC in those with comorbid AD (n=32). **F**, Plasma TDP-43 levels were significantly elevated in individuals with hippocampal sclerosis (P = 0.048). Boxplots show median values with interquartile ranges, and ROC curves visualize sensitivity versus 1-specificity for classification.

To our knowledge, this is the first time that significant associations of plasma TDP-43 and pTDP-43 with autopsy-confirmed advanced LATE-NC are demonstrated, particularly in individuals with comorbid AD. Plasma TDP-43 was also elevated in HS, a pathologic finding closely related to advanced LATE-NC, showing internally consistent results. Notably, plasma pTDP-43, which was reported to be elevated in ALS [4], did not perform as well as total TDP-43 in detecting advanced LATE-NC. This discrepancy might be attributed to the lower concentration of pTDP-43, which is closer to the detection threshold of current assays [6]. Similarly, better biomarker performance observed in the AD subgroup might be driven by higher LATE-NC burden in AD. Intriguingly, plasma TDP-43 and pTDP-43 were inversely correlated with tau, which we could not further examine due to limited sample size. Nevertheless, tau or other pathologies did not confound the LATE-NC plasma biomarker association.

Our results highlight the promise of plasma-based high-sensitivity TDP-43 assays for detecting LATE-NC, particularly in those with comorbid AD. Future larger studies are necessary to validate these findings and to confirm the utility of plasma TDP-43 in AD/LATE clinical trials or clinical practice (e.g., impact on AD disease-modifying treatments).

## Data Availability

All data analyzed in this manuscript can be requested at the RADC Resource Sharing Hub at https://www.radc.rush.edu. All data used in this study are individual-level human data that require the investigators to sign a data use agreement to ensure human subject protection.

## Ethics approval and consent to participate

All study protocols and procedures were approved by the Institutional Review Board at Rush University Medical Center, and all participants signed written informed consent and the Anatomical Gift Act.

## Author Contributions

J.W., T.L.Y.-P., and H.-S.Y. conceptualized and designed the study. J.A.S., D.A.B., N.T.S., and T.L.Y.-P acquired the data. J.W. and H.-S.Y. performed statistical analyses, generated figures, and drafted the manuscript. All authors interpreted the results, revised the manuscript for important intellectual content, and approved the manuscript.

## References

[1] Nelson PT, Dickson DW, Trojanowski JQ, et al. Limbic-predominant age-related TDP-43 encephalopathy (LATE): consensus working group report. Brain. Jun 1 2019;142(6):1503–1527. doi:10.1093/brain/awz099

[2] Wolk DA, Nelson PT, Apostolova L, et al. Clinical criteria for limbic-predominant age-related TDP-43 encephalopathy. Alzheimers Dement. Jan 2025;21(1):e14202. doi:10.1002/alz.14202

[3] Meneses A, Koga S, O’Leary J, Dickson DW, Bu G, Zhao N. TDP-43 Pathology in Alzheimer’s Disease. Mol Neurodegener. Dec 20 2021;16(1):84. doi:10.1186/s13024-021-00503-x

[4] Thomas EV, Han C, Kim WJ, et al. ALS plasma biomarkers reveal neurofilament and pTau correlate with disease onset and progression. Ann Clin Transl Neurol. Feb 6 2025; doi: 10.1002/acn3.70001

[5] Winston CN, Sukreet S, Lynch H, et al. Evaluation of blood-based, extracellular vesicles as biomarkers for aging-related TDP-43 pathology. Alzheimers Dement (Amst). 2022;14(1):e12365. doi:10.1002/dad2.12365

[6] Zeng X, Lafferty TK, Sehrawat A, et al. Multi-analyte proteomic analysis identifies blood-based neuroinflammation, cerebrovascular and synaptic biomarkers in preclinical Alzheimer’s disease. Mol Neurodegener. Oct 10 2024;19(1):68. doi:10.1186/s13024-024-00753-5

[7] Bennett DA, Buchman AS, Boyle PA, Barnes LL, Wilson RS, Schneider JA. Religious Orders Study and Rush Memory and Aging Project. J Alzheimers Dis. 2018;64(S1):S161–S189. doi:10.3233/JAD-179939

[8] Lish, A. M. et al. CLU alleviates Alzheimer’s disease-relevant processes by modulating astrocyte reactivity and microglia-dependent synaptic density. Neuron, doi:10.1016/j.neuron.2025.03.034 (2025).

