## Supplementary materials for "Plasma TDP-43 is a potential biomarker for advanced limbic-predominant age-related TDP-43 encephalopathy neuropathologic change"

**Supplementary Table. Characteristics of study participants.**

| | All | AD Dementia | Control |
|---|---|---|---|
| N | 50 | 32 | 18 |
| Age at blood draw (mean±SD) | 89.9±5.2 | 91.3±5.2 | 87.5±4.3 |
| Age at death (mean, SD) | 93.8±5.3 | 95.8±4.7 | 90.2±4.2 |
| Female, n (%) | 38 (76%) | 23 (72%) | 12 (67%) |
| Pathologic diagnosis of AD | 32 (64%) | 32 (100%) | 0 (0%) |
| LATE-NC stage, n (%) | | | |
| 0, none | 15 (30%) | 3 (9%) | 12 (67%) |
| 1, amygdala | 12 (24%) | 9 (28%) | 3 (17%) |
| 2, limbic | 8 (16%) | 5 (16%) | 3 (17%) |
| 3, neocortex | 15 (30%) | 15 (47%) | 0 (0%) |
| Hippocampal sclerosis | 4 (8%) | 4 (12.5%) | 0 (0%) |
| Any Lewy body pathology, n (%) | 18 (36%) | 14 (44%) | 4 (22%) |

Demographic characteristics and pathology assessment information of the subjects from ROSMAP cohort. AD, Alzheimer's disease; SD, standard deviation; LATE-NC, limbic-predominant age-related TDP-43 encephalopathy neuropathologic change.
